# Factors Associated with Acute Kidney Injury in COVID-19 Hospitalized Patients in Central Java, Indonesia

**DOI:** 10.1101/2024.07.30.24311116

**Authors:** Dwi Lestari Partiningrum, Melissa Angela Chionardes, Nurul Hasanah Yusri, Indra Adhim Karunia Aji, Jonathan Christianto Subagya, Aldrich Kurniawan Liemarto

**Affiliations:** Diponegoro University, Faculty of Medicine, Department of Internal Medicine, Division of Nephrology and Hypertension, Semarang, Central Java, Indonesia; Columbia Asia Hospital, Department of Internal Medicine, Semarang, Central Java, Indonesia; Columbia Asia Hospital, Semarang, Central Java, Indonesia; Diponegoro University, Faculty of Medicine, Master Program of Biomedical Science, Semarang, Central Java, Indonesia; UNIMUS Hospital, Semarang, Central Java, Indonesia; Cilincing Regional Public Hospital, North Jakarta, Jakarta, Indonesia

**Keywords:** Acute kidney injury, COVID-19, risk factors, Central Java, Indonesia

## Abstract

**Backgroud and Objective:** Acute kidney injury (AKI) is a significant complication of COVID-19 infection, with varied incidence rates globally. COVID-19 has exacerbated AKI cases, with a significant portion of patients experiencing kidney damage. This study investigates the prevalence and risk factors associated with AKI among COVID-19 patients in Semarang, Central Java, Indonesia.

**Methods:** Data from 364 hospitalized COVID-19 patients in a hospital in Semarang between March 2020 and September 2021 were analyzed. Statistical analysis using chi-square and logistic regression examined the relationship between AKI and its determinants, with p≤0.05 considered significant.

**Results:** The majority of patients were male, most had no prior medical conditions. Analysis indicated links between AKI and various factors like several physical and supportive examination results. Few comorbidities were found to increase the risk of AKI, followed also by abnormal vital signs except blood pressure, several elevated level of laboratory results, and radiologic pneumonia finding.

**Conclusion:** COVID-19 may harm the kidneys causing AKI. This study highlights the importance of history taking, examination, and laboratory monitoring to detect AKI in COVID-19 patients.

## Introduction

Acute kidney injury (AKI) represents a kidney disorder associated with notably high morbidity and mortality rates, impacting up to 13 million individuals globally and contributing to 1.7 million deaths. Reports suggest that 80% of AKI cases worldwide originate in developing countries, with Southeast Asia exhibiting the highest prevalence at 31%.^1,2^ AKI is characterized by a sudden onset of diminished kidney function, typically manifested by a decline in serum creatinine levels or urine output within a span of hours to days.^3^ Etiologies of AKI are diverse, including ischemia, infection, exposure to nephrotoxic agents, among others. Particularly in developing nations, AKI often intersects with endemic infectious diseases prevalent in those regions. Microbial infections have the potential to directly impair kidney parenchymal tissue or precipitate sepsis, which can inflict damage on multiple organ systems.^1,4,5^

As of 2023, COVID-19 has reportedly affected 6.8 million individuals in Indonesia, resulting in over 161,900 deaths, with the potential for further increases. The country’s geographical landscape, comprising numerous islands, along with varied socioeconomic statuses, presents challenges in controlling disease transmission.^6^ The Special Capital Region of Jakarta, West Java, and Central Java are the three provinces in Indonesia with the highest number of COVID-19 cases.^7^ While some individuals infected with COVID-19 may only exhibit mild symptoms such as fever, cough, and myalgia, others may experience more severe manifestations, including acute respiratory distress syndrome. SARS-CoV-2, the virus responsible for COVID-19, is a positive-sense single-stranded RNA virus belonging to the Coronaviridae family. SARS-CoV-2 possesses the ability to mutate and generate new variants through antigenic evolution, enabling the virus to evade host immunity, even among vaccinated individuals. The continuous evolution of the virus presents challenges in efforts to eradicate it.

Part of the Indonesian population is still hesitant to receive the COVID-19 booster vaccinations. Booster vaccine acceptance in Indonesia remains below the national vaccination coverage target of 69.2%. They do not seek booster vaccinations because they believe that innate immunity is sufficient to prevent COVID-19 and that the first vaccination they obtained is ineffective against COVID-19.^8^ Antibody immunity conferred by primary immunization would wane over time if not re-induced with booster vaccine. Hence, COVID-19 transmission in Indonesia cannot yet be completely avoided.^9^ Furthermore, the vast majority of Indonesians receive an initial vaccination in the form of an inactivated vaccine. Inactivated vaccinations are less effective than mRNA vaccines in preventing severe COVID-19 and mortality.^10^

COVID-19 is also capable of causing multi-organ damage, with the kidneys being particularly susceptible, leading to AKI. A meta-analysis revealed that 5.5% of Chinese COVID-19 inpatients and 28.6% of American and European COVID-19 inpatients experienced AKI, with a mortality risk ratio of 4.6.^11^ The main pathogenesis of COVID-19 is attributed to SARS-CoV-2’s ability to bind to all angiotensin-converting enzyme 2 (ACE2) receptors in humans.^12^ These receptors are present in kidney proximal tubule cells and podocytes. SARS-CoV-2 can cause direct damage by fusing with cells expressing ACE2 receptors. Once inside the cell’s cytoplasm, SARS-CoV-2 RNA activates inflammatory signaling pathways. Additionally, AKI associated with COVID-19 can be induced by mechanisms such as cytokine storm, complement activation, hypercoagulability, and hemodynamic instability.^13,14^ Several risk factors have been identified as contributing to AKI development in COVID-19 patients. These include diabetes mellitus, hypertension, chronic kidney disease, hyperlipidemia, cardiovascular disease, older age, smoking, immunocompromised conditions, high body mass index, ethnicity, and abnormalities in specific laboratory results.^15^

The COVID-19 pandemic in Indonesia has transitioned to endemic status, signifying a shift in the ongoing response efforts. However, vigilance remains paramount. Continuous discourse surrounding COVID-19-associated AKI persists, given the myriad complexities unearthed and the evolving understanding of the disease. The objective of this study is to delve deeper into the relationship between various risk factors that could contribute to AKI in COVID-19 patients, with a particular focus on hospital patients in Semarang city. Semarang city stands as one of the primary epicentres for COVID-19 cases in Indonesia, making it a crucial locale for this investigation.

## Methods

This investigation adopted a cross-sectional design and transpired between March 2020 and September 2021 within a hospital located in Semarang, Central Java, Indonesia. The participant pool consisted of 364 subjects recruited during their hospital admissions, with exclusion criteria applied to individuals lacking comprehensive laboratory test results. This study utilizes data including symptoms, laboratory results, and radiologic findings extracted from electronic medical records that have obtained approval from the ethical committee, with number 0250/DIR/KEH/RSKBCAS/2023.

The determination of Body Mass Index (BMI) involved the measurement of body weight and height using a stature meter and weight scale. BMI was classified according to the WHO Asia Pacific category. Blood pressure was assessed utilizing a sphygmomanometer and subsequently classified into normal or hypertension. Vital signs, excluding blood pressure, were categorized as either normal or elevated. Oxygen saturation levels were distinguished as either normal or declined. Additional laboratory assessments were conducted, and results were categorized as normal or abnormal. Acute Kidney Injury (AKI) was established as serum creatinine levels equal to or exceeding 1.2 mg/dL.

Statistical analyses employed bivariate chi-square and multivariate logistic regression methods to scrutinize the relationship between AKI and its determinant factors. A p-value equal to or less than 0.05 (≤0.05) was deemed statistically significant.

## Results

During March 2020 to September 2021, 1084 COVID-19 patients were recorded, but only 364 subjects met the inclusion criteria. There was a higher prevalence of male patients. The majority of patients lacked a medical history of ailments, as shown in Table 1. Based on the bivariate analysis, relationships were observed between AKI and variables such as age, comorbidities of diabetes, hypertension, chronic kidney disease, history of hemodialysis, heart diseases, admission to intensive care unit, use of ventilators and vasopressor drugs. From physical and supportive examinations, vital signs except blood pressure were correlated to AKI, followed by erythrocyte count, thrombocyte count, lymphocyte, hemoglobin, hematocrit, ureum, sodium, serum glutamate-pyruvate transaminase (SGPT), and radiologic pneumonia finding.

**Table 1.** Baseline Characteristics.

| Characteristics |  | n | % |
| --- | --- | --- | --- |
| Age | <25 | 5 | 1.4 |
|  | 25-44 | 96 | 26.4 |
|  | 45-59 | 145 | 39.8 |
|  | 60-74 | 96 | 26.4 |
|  | 75-90 | 21 | 5.8 |
|  | >90 | 1 | 0.3 |
| Gender | Male | 217 | 59.6 |
|  | Female | 147 | 40.4 |
| History of Diabetes | Yes | 103 | 28.3 |
| History of Hypertension | Yes | 114 | 31.3 |
| History of Chronic Kidney Disease | Yes | 6 | 1.6 |
| History of Hemodialysis | Yes | 3 | 0.8 |
| History of Stroke | Yes | 8 | 2.2 |
| History of Heart Diseases | Yes | 25 | 6.9 |
| History of Hyperlipidemia | Yes | 8 | 2.2 |
| History of Liver Diseases | Yes | 3 | 0.8 |
| History of Lung Diseases | Yes | 13 | 3.6 |
| History of Thyroid Diseases | Yes | 2 | 0.5 |
| History of Malignancy | Yes | 3 | 0.8 |
| Fever | Yes | 265 | 72.8 |
| Diarrhea | Yes | 28 | 7.7 |
| Smoking | Yes | 1 | 0.3 |
| ICU | Yes | 74 | 20.3 |
| HFNC | Yes | 6 | 1.6 |
| NIV | Yes | 32 | 8.8 |
| Invasive Ventilator | Yes | 20 | 5.5 |
| Azithromycin | Yes | 81 | 22.3 |
| Hydro Chloroquinolone | Yes | 3 | 0.8 |
| ACEi/ARB | Yes | 73 | 20.1 |
| Vasopressor | Yes | 34 | 9.3 |
| Body Mass Index | Abnnormal | 295 | 81.0 |
| Blood Pressure | Elevated | 316 | 86.8 |
| Heart Rate | Tachycardia | 80 | 22.0 |
| Respiratory Rate | Tachypnea | 103 | 28.3 |
| Temperature | Elevated | 62 | 17.0 |
| SpO2 | Low | 163 | 44.8 |
| Erythrocyte | Decreased | 56 | 15.4 |
| Leukocyte | Abnormal | 85 | 23.4 |
| Thrombocyte | Low | 64 | 17.6 |
| Hemoglobin | Normal | 56 | 15.44 |
| Hematocrite | Low | 130 | 35.7 |
| MCV | Abnormal | 58 | 15.9 |
| MCH | Abnormal | 115 | 31.6 |
| MCHC | Abnormal | 135 | 37.1 |
| RDW | Elevated | 53 | 14.6 |
| Eosinophil | Elevated | 13 | 3.6 |
| Basophil | Elevated | 3 | 0.8 |
| Neutrophil | Abnormal | 241 | 66.2 |
| Lymphocyte | Abnormal | 313 | 86.0 |
| Monocyte | Elevated | 260 | 71.4 |
| Ureum | Elevated | 235 | 64.6 |
| Creatinine | Elevated | 84 | 23.1 |
| Natrium Serum | Abnormal | 168 | 46.2 |
| Kalium Serum | Abnormal | 90 | 24.7 |
| Chloride Serum | Abnormal | 131 | 36.0 |
| SGOT | Elevated | 156 | 42.9 |
| SGPT | Elevated | 116 | 31.9 |
| Glucose | Elevated | 84 | 23.1 |
| C-Reactive Protein | Elevated | 358 | 98.4 |
| D-dimer | Elevated | 364 | 100 |

ICU = Intensive Care Unit, HFNC = High Flow Nasal Cannula, NIV = Non-Invasive Ventilator, ACEi = Angiotensin-Converting Enzyme Inhibitor, ARB = Angiotensin Receptor Blocker, SpO2 = Saturation of Oxygen, MCV = Mean Corpuscular Volume, MCH = Mean Corpuscular Hemoglobin, MCHC = Mean Corpuscular Hemoglobin Concentration, RDW = Red cell Distribution Width, SGOT = Serum Glutamic Oxaloacetic Transaminase, SGPT = Serum Glutamic Pyruvate Transaminase

Multivariate analysis revealed that comorbidities acted as risk factors against AKI, such as history of diabetes, hypertension, and heart diseases. Through supportive examinations, the foremost risk factors included elevated urea levels, followed by serum glutamate-oxaloacetate transaminase (SGOT), thrombocyte count, and serum chloride, as shown in table 3.

**Table 2.** Relationship between Comorbidities, Physical, and Supportive Examinations with AKI in COVID-19 Patients.

| **Patients Characteristics** | **No AKI (%)** | **AKI (%)** | **p Value** |
| --- | --- | --- | --- |
| **Medical History** |  |  |  |
| Old age | 74 (62.7) | 44 (37.3) | <0.001 |
| **Comorbidities** |  |  |  |
| Diabetes | 66 (23.6) | 37 (44.0) | <0.001 |
| Hypertension | 72 (25.7) | 41 (50.0) | <0.001 |
| CKD | 0 (0) | 6 (7.1) | <0.001 |
| Haemodialysis | 0 (0) | 3 (3.6) | 0.012 |
| Stroke | 5 (1.8) | 3 (3.6) | 0.393 |
| Heart Diseases | 14 (5.0) | 11 (13.1) | 0.010 |
| Hyperlipidaemia | 4 (1.4) | 4 (4.8) | 0.087 |
| Liver Diseases | 2 (0.7) | 1 (1.2) | 0.546 |
| Lung Diseases | 11 (3.9) | 2 (2.4) | 0.740 |
| Thyroid Diseases | 2 (0.7) | 0 (0) | 1.000 |
| Malignancy | 1 (0.4) | 2 (2.4) | 0.134 |
| History of Fever | 205 (73.2) | 60 (71.4) | 0.747 |
| History of Diarrhea | 23 (8.2) | 5 (6.0) | 0.495 |
| History of Smoking | 0 (0) | 1 (1.2) | 0.231 |
| Intensive Care Unit | 47 (16.8) | 27 (32.1) | 0.002 |
| Use of High Flow Nasal Cannula | 4 (1.4) | 2 (2.4) | 0.625 |
| Use of Non-invasive Ventilator | 20 (7.1) | 12 (14.3) | 0.043 |
| Use of Invasive Ventilator | 9 (3.2) | 11 (13.1) | 0.001 |
| **Drugs Use** |  |  |  |
| Azithromycin | 62 (22.1) | 19 (22.6) | 0.927 |
| Hydroxychloroquine | 2 (0.7) | 1 (1.2) | 0.546 |
| ACEi/ARB | 49 (17.5) | 24 (28.6) | 0.026 |
| Vasopressor | 16 (5.7) | 18 (21.4) | <0.001 |

| **Physical and Supportive Examination** |  |  |  |
| --- | --- | --- | --- |
| Body Mass Index | 280 (100) | 83 (98.8) | 0.231 |
| Blood Pressure | 242 (86.4) | 74 (88.1) | 0.692 |
| Heart Rate | 54 (19.3) | 26 (31.0) | 0.024 |
| Respiratory Rate | 71 (25.4) | 32 (38.1) | 0.023 |
| Temperature | 41 (14.6) | 21 (25.0) | 0.027 |
| Oxygen Saturation | 112 (40.0) | 51 (60.7) | <0.001 |
| Erythrocyte | 33 (11.8) | 23 (27.4) | <0.001 |
| Leukocyte | 64 (22.9) | 21 (25.0) | 0.684 |
| Thrombocyte | 37 (13.2) | 27 (32.1) | <0.001 |
| MCV | 45 (16.1) | 13 (15.5) | 0.896 |
| MCH | 93 (33.2) | 22 (26.2) | 0.225 |
| MCHC | 106 (37.9) | 29 (34.5) | 0.579 |
| RDW | 39 (13.9) | 14 (16.7) | 0.533 |
| Neutrophil | 178 (63.6) | 63 (75.0) | 0.052 |
| Lymphocyte | 234 (83.6) | 79 (94.0) | 0.015 |
| Monocyte | 205 (73.2) | 55 (65.5) | 0.169 |
| Eosinophil | 12 (4.3) | 1 (1.2) | 0.313 |
| Basophil | 3 (1.1) | 0 (0) | 1.000 |
| Hemoglobin | 30 (10.7) | 26 (31.0) | <0.001 |
| Haematocrit | 85 (30.4) | 45 (53.6) | <0.001 |
| Ureum | 153 (54.6) | 82 (97.6) | <0.001 |
| Natrium | 111 (39.6) | 57 (67.9) | <0.001 |
| Kalium | 65 (23.2) | 25 (29.8) | 0.222 |
| Chloride | 101 (36.1) | 30 (35.7) | 0.952 |
| SGOT | 113 (40.4) | 43 (51.2) | 0.078 |
| SGPT | 110 (35.7) | 16 (19.0) | 0.004 |
| Glucose | 59 (21.1) | 25 (29.8) | 0.097 |
| CRP | 274 (97.9) | 84 (100) | 0.343 |
| D-dimer | 280 (100) | 84 (100) | Constant |
| Radiologic Pneumonia Finding | 248 (88.6) | 82 (97.6) | 0.012 |

CKD = Chronic Kidney Disease, ACEi = Angiotensin-Converting Enzyme Inhibitor, ARB = Angiotensin Receptor Blocker, SpO<sub>2</sub> = Saturation of Oxygen, MCV = Mean Corpuscular Volume, MCH = Mean Corpuscular Hemoglobin, MCHC = Mean Corpuscular Hemoglobin Concentration, RDW = Red cell Distribution Width, SGOT = Serum Glutamic Oxaloacetic Transaminase, SGPT = Serum Glutamic Pyruvate Transaminase

**Table 3.** Risk Factors of AKI in COVID-19 Patients.

| Comorbidities | p | OR | 95% CI |  |
| --- | --- | --- | --- | --- |
|  |  |  | Lower | Upper |
| History of Diabetes | 0.017 | 2.065 | 1.140 | 3.740 |
| History of Hypertension | 0.008 | 2.442 | 1.261 | 4.730 |
| History of Heart Diseases | 0.013 | 3.678 | 1.317 | 10.271 |
| Thrombocyte | 0.001 | 3.741 | 1.667 | 8.392 |
| Ureum | <0.001 | 24.440 | 5.477 | 109.054 |
| Chloride | 0.002 | 2.724 | 1.424 | 5.213 |
| SGOT | <0.001 | 5.013 | 2.175 | 11.554 |

## Discussion

Incidence of Acute Kidney Injury (AKI) documented among individuals with COVID-19 has varied, spanning from 0.5% to 36.6%.^16^ Meanwhile, in a study conducted in Bahrain, 47.6% of hospitalized patients diagnosed with COVID-19 experienced the development of AKI.^17^ The destructive effects of SARS-CoV-2 on podocytes and proximal tubule cells may contribute to the development of COVID-19 AKI. Viremia could cause endothelial injury, promoting vasoconstriction, hypercoagulability, and activation of macrophages, ultimately leading to the formation of microthrombi and injury to the renal microvasculature. Moreover, the loss of functional nephrons may augment the development of renal fibrosis. Decreased kidney perfusion was also observed in COVID-19 pneumonia as result of right ventricular failure.^18^

This study identified a correlation between history of comorbidity, the utilization of mechanical ventilators, intensive care unit (ICU) admission, and specific medications with AKI. A meta-analysis findings indicated a significant association between various comorbidities such as diabetes, ischemic heart disease, heart failure, hypertension, and chronic kidney disease with AKI. Additionally, the utilization of mechanical ventilation was also significantly associated with AKI in COVID-19 patients. Indeed, there is a notable association between oxidative stress, atherosclerosis, and endothelial dysfunction in comorbid diseases, which may serve as the underlying mechanism for their relationship with AKI.^19,20^ In this study, the use of vasoactive drugs were associated with AKI in COVID-19, as seen in the study by de Almeida et al. Patients who undergo mechanical ventilation, require admission to the ICU, and receive vasopressor drugs may be at higher risk of experiencing severe AKI.^21^

Several laboratory profiles exhibited significant disparities between individuals who developed between AKI and non-AKI groups. Patients who developed AKI demonstrated notably elevated levels of ureum, SGPT, and D-dimer, while displaying decreased levels of erythrocytes, thrombocytes, hemoglobin, hematocrit, and lymphocytes. Consistent with earlier investigations conducted by Naser et al in 2021, which revealed comparable laboratory findings, albeit with the exception of erythrocyte values due to the absence of data in their study.^17^

Laboratory parameters examined in a study conducted by Xu et al in 2022 in China demonstrated largely comparable outcomes, although notable distinctions were observed in platelet, leukocyte, neutrophil, lymphocyte, and urea values between the AKI and non-AKI groups.^22^ Nevertheless, sodium levels did not exhibit variance between these groups. Notably, potassium levels were found to be elevated in this study. The observation regarding thrombocyte and D-dimer may suggest that COVID-19 patients with AKI experienced impairment in coagulation.^23^ Meanwhile ureum level showed that kidney function in COVID-19 patients with AKI were worse. A meta-analysis also showed that elevated D-dimer levels, alongside diminished platelet count, and hemoglobin levels were correlated with severe COVID-19.^24^

The history of hypertension and diabetes mellitus in our study is an independent risk factor for the development of AKI in COVID-19 patients. These results are consistent with the results of a systematic review by Sabaghian et al., where hypertension and diabetes mellitus (DM) are comorbid diseases that are commonly found in COVID-19 patients with AKI at 61.4% and 40%, respectively.^15^ Vascular endothelial dysfunction is known to be caused by COVID-19, hypertension, and diabetes mellitus.^25^ The cytokine storm caused by SARS-CoV-2 leads to systemic endothelial dysfunction. A high amount of TNF-α will cause a decrease in levels of endothelial NO synthase (eNOS), which shortens the half-life of nitric oxide (NO), one of the most potent vasodilators, while IL-1β suppresses endothelial proliferation.^26^ Hypertension can cause endothelial dysfunction through impaired shear stress of blood vessels and activation of the renin-angiotensin system.^27^ Shear stress disrupting blood flow triggers mechanoreceptors, which in turn suppress the production of NO. Angiotensin-II is also known to aggravate oxidative stress and inflammation by inducing a higher production of aldosterone and NADPH oxidase, both of which are generated constantly in hypertension patients.^27-29^ Diabetic patients also have decreased NO levels. Patients with diabetes mellitus (DM) express less insulin receptor substrate 2 (IRS-2), including less of it in the endothelium, causing insulin resistance. Insulin phosphorylates IRS-2 via binding to its receptor. The PI3-K/Akt signaling pathway, which involves the role of Akt in regulating NO generation and endothelial proliferation, is activated as a result of this phosphorylation. Reduced NO production will lead to endothelial damage through vasoconstriction, disturbance of laminar blood flow, and ROS accumulation. When endothelium becomes damaged, it releases a variety of pro-inflammatory cytokines and chemokines that lead to the recruitment of leukocytes, ensuing local inflammation, and ending in massive endothelial damage.^30,31^ Endothelial dysfunction that occurs in the kidney can trigger AKI. The glomerulus and peritubular capillaries have a fenestrated endothelium, which plays a role in the processes of filtration, reabsorption, and secretion. Damage to the endothelial structure due to endothelial dysfunction causes hyperpermeability, so that kidney function becomes impaired.^32^

Additionally, our findings indicate that heart disease is also significantly associated with the AKI in COVID-19 patients. AKI risk is higher in COVID-19 patients with heart failure and ischemic heart disease, according to a systematic study by Hidayat et al.^19^ A cohort study by Kolhe et al. also found that COVID-19 patients with congestive heart failure had an odds ratio of 1.72 for the occurrence of AKI.^33^ That condition is caused by a decrease in cardiac output or increased renal venous pressure.^34^ The majority of our research population with heart disease also had a history of hypertension or DM, suggesting that heart disease may have developed as a consequence of these two conditions. As a result, heart disease was also an independent risk factor in our analysis.

Other autonomous risk factors associated with AKI in COVID-19 patients comprised thrombocyte, elevated urea levels, chloride, and SGOT. The attachment of SARS-CoV-2 to the ACE2 receptor reduces ACE2 function and increases angiotensin II, resulting in oxidative stress and endothelial dysfunction, which triggers platelet hyperactivity. Furthermore, thrombopoiesis will increase due to hypoxia, which is induced by a decrease in oxygen saturation and massive platelet apoptosis in COVID-19 patients. This will result in microvascular thrombosis in multiple organs, including the kidneys.^35,36^ Chloride can indeed induce vasoconstriction, particularly in renal vessels, highlighting a specific effect on kidney function, thus leading to AKI.^37^ In the study by Khruleva et al., elevated SGOT was a common significant factor associated with phenotypes of AKI. Potential contributing factors may encompass ischemic damage, damage induced by systemic inflammation akin to sepsis, and coagulation abnormalities involving both microvascular and macrovascular thrombosis.^38^

The kidneys are susceptible to damage during COVID-19 infection, leading to acute kidney injury (AKI). This study observed a 23.3% incidence of increased creatinine among patients with COVID-19. History of several chronic diseases such as heart diseases, hypertension, and diabetes were risk factors of AKI in COVID-19. Laboratory and radiological investigations yielded variable results, with urea emerging as the most significant risk factor for AKI. This study underscores the importance of monitoring various laboratory parameters that may contribute to AKI in COVID-19 patients. To the author’s knowledge, this is the first study examining risk factors for AKI in Semarang, Central Java. Future research could expand by involving multiple centers to gather data, thus enhancing the generalizability of findings within Central Java and Indonesia.

## Data Availability

All data produced in the present work are contained in the manuscript

## Notes

**Name of the funding agency** This study was not supported by any sponsor or funder.

**Conflict of interest** All authors declare that there is no conflict of interest.

### Competing Interest Statement

The authors have declared no competing interest.

### Funding Statement

This study did not receive any funding

### Author Declarations

Ethics committee of Columbia Asia Hospital gave ethical approval for this work

